# Changes in monocyte subsets can predict the risk of AAA and are surrogate markers for AAA morphology in patients with late-stage disease

**DOI:** 10.1101/2024.11.18.24317518

**Authors:** Bianca Hamann, Anna Klimova, Marvin Kapalla, David M. Poitz, Frieda Frank, Henning Morawietz, Christian Reeps, Anja Hofmann

**Affiliations:** Division of Vascular and Endovascular Surgery, Department of Visceral, Thoracic and Vascular Surgery, University Hospital and Medical Faculty Carl Gustav Carus, TUD Dresden University of Technology, Germany; Institute for Medical Informatics and Biometry, Faculty of Medicine, TUD Dresden University of Technology, Germany; Institute for Clinical Chemistry and Laboratory Medicine; University Hospital and Medical Faculty Carl Gustav Carus, TUD Dresden University of Technology, Germany; Division of Vascular Endothelium and Microcirculation, Department of Medicine III, University Hospital and Medical Faculty Carl Gustav Carus, TUD Dresden University of Technology, Germany

## Abstract

**Background:** Monocytes play a pivotal role in pathology of abdominal aortic aneurysm (AAA) and can display an immunophenotypic heterogeneity with functionally distinct subpopulations. Alterations in monocyte subsets have been described in CVD, and some are associated with cardiovascular risk, but their profile in AAA is poorly understood.

**Aim:** We aimed to comprehensively define associations of circulating monocyte phenotypes with AAA risk and AAA morphology.

**Methods:** Monocyte subsets (CD14++CD16−; CD14++/CD16+; CD14+/CD16++) were analyzed in a prospective, observational study in patients with AAA (n=34) and varicose veins (n=34) by using flow cytometry.

**Results:** Classical monocytes were 1.6-fold lower (*P*=0.001) in AAA, while intermediate and non-classical monocytes were 1.8 (*P*=0.019) and 1.9-fold (*P*=0.025) higher in AAA, respectively. The differences remained significant after adjusting for age, sex and peripheral artery disease. A lower proportion of classical monocytes (HR: 0.73, *P*=0.002) and increases in intermediate (HR: 1.41, *P*=0.006) and non-classical monocytes (HR: 1.54, *P*=0.030) were associated with a higher risk of AAA. Non-classical monocytes showed an inverse correlation with AAA diameter (Pearson correlation =-0.64, *P*=0.001) and AAA volume (Pearson correlation =-0.50, *P=*0.003).

**Conclusion:** The present study revealed age- and sex-independent shifts in monocytes, all of which were associated with risk of AAA disease. Non-classical monocytes were inversely correlated with AAA diameter and volume and thus may be surrogate markers for AAA morphology.

**What’s new?:**

- Classical monocytes are lower in patients with late-stage AAA.
- Non-classical monocytes showed the strongest increase in AAA disease.
- A reduction in classical monocytes is associated with increased risk of AAA.
- An increase in non-classical and intermediate monocytes is associated with an increased risk of AAA.
- Lowering in non-classical monocytes may be a surrogate marker for AAA morphology, particlarly AAA volume.
- Intermediate monocytes showed a positive correlation with the thickness of the intraluminal thrombus.

**What are the clinical implications?:**

- The increase in non-classical monocytes could be a novel surrogate marker for AAA volume, which could be useful when AAA diameter is insufficient or to monitor a saccular AAA, as there is a weaker correlation between diameter and risk of rupture in this type of AAA.
- The decrease in classical monocytes could be a useful surrogate marker for AAA volume and provide additional information on AAA diameter.
- Monocyte shifts and their association with AAA disease may be relevant for the diagnosis of AAA and should be verified in larger cohorts.
- The mechanisms behind the decrease in classical monocytes and the increase in intermediate and non-classical subsets should be investigated in further *in vitro* studies as they offer therapeutic potential.

**Graphical Abstract:** 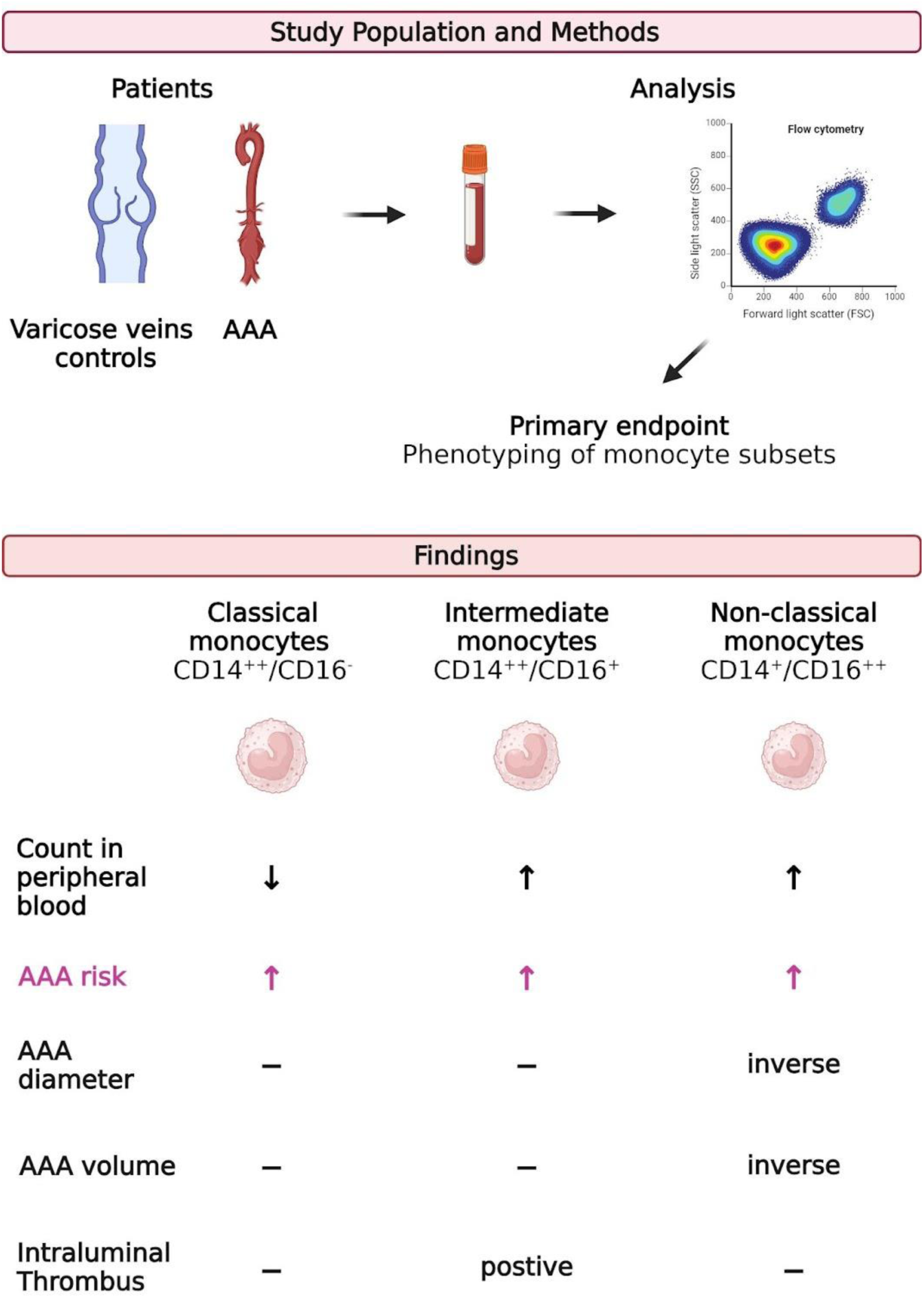

## Introduction

Abdominal aortic aneurysm (AAA) is a life-threatening condition with high mortality rates. Main pathological features are inflammation with increased local cytokine production, extracellular matrix degradation, oxidative stress and the loss of smooth muscle cells.^1^ The only effective treatment is surgical elimination of the AAA. Pharmacological treatments are still lacking, emphasizing the need to define the pathomechanisms that contribute to the initiation and progression of the disease. Inflammatory cells in AAA comprise monocytes, macrophages, neutrophils, mast cells, natural killer (NK) cells, T and B cells and dendritic cells.^2^ Monocytes are recruited at sites of inflammation where they differentiate into macrophages.^3^ Monocytes exist in subsets, which differ in morphology, function and cell surface marker (e.g. chemokine receptors) expression. Three distinct subsets were distinguished in humans based on the expression of the the LPS-receptor CD14 and the Fcγ receptor CD16: classical (CD14++CD16−), non-classical (CD14+CD16++) and intermediate (CD14++CD16+) monocytes.^4^ Classical monocytes (CD14++CD16−) secrete large amounts of pro-inflammatory cytokines and promote angiogenesis, wound healing and coagulation, demonstrating their importance in tissue repair.^3^ In contrast, CD16-expressing cells may secrete pro-inflammatory cytokines, have a high antigen-presenting capacity and the ability to migrate into subendothelial spaces and atherosclerotic lesions.^4^ These cells include intermediate monocytes (CD14++CD16+), that secrete pro-inflammatory cytokines and accumulate in chronic inflammatory diseases.^3,5,6^ Intermediate subsets are predictive for cardiovascular disease^4^ and are associated, for example, with susceptibility to plaque vulnerability^3^. CD16+ subsets are elevated in patients with AAA^7^ and a recently published study showed that only intermediate monocytes (CD14++CD16+) were elevated in AAA compared to healthy individuals and patients with PAD^8^. Interestingly, CD14++CD16+ and D-dimer proved to be suitable diagnostic markers for AAA while elevated concentrations of these subsets were able to predict rapid growing AAA.^8^

Non-classical monocytes have low expression of CD14 (CD14+) and secrete pro- and anti-inflammatory cytokines. These monocytes have phagocytic properties to remove cell debris and are important for patrolling the vascular endothelium and controlling vascular integrity. Most likely, they are involved in the resolution of inflammation.^3^ However, it should be noted that a CD14-low subset may promote neovascularization and destabilization of lesions, while their accumulation in ischemic tissue may have beneficial effects.^4^

Although an increase in intermediate subsets has been demonstrated in AAA, it is still under debate whether the three subgroups correlate with AAA diameter, AAA volume, and thickness of the intraluminal thrombus as the main morphologic features of AAA and could be useful surrogate markers. In addition, there is no information whether any of these monocyte subsets can predict end-stage AAA, when most patients with AAA are diagnosed. Here, we aimed to comprehensively define associations of circulating monocyte phenotypes with AAA risk and AAA morphology.

## Materials and Methods

### Study Design

Monocyte subpopulations were determined in an observational study in electively treated AAA patients and patients with varicose veins who served as controls. Due to the observational nature of the study, no sample size or power calculation was performed, according to the STROBE guidelines.^9^ Patients were treated in the Department of Visceral, Thoracic and Vascular Surgery between December 2020 and June 2024. The diagnostic criteria for varicose patients included venous reflux assessed by duplex ultrasound and signs of chronic venous insufficiency according to the CEAP classification.^10^ Inclusion criteria were age over 50 years and no known history of arterial disease, T2D and infectious diseases. Patients with thrombophlebitis were excluded. Inclusion criteria for AAA patients were a diameter of the infrarenal aorta of more than 40 mm, a rapidly growing AAA with more than 10 mm progression per year or a symptomatic AAA. The study was approved by the ethics committee of the Technische Universität Dresden (EK 151042017). Informed consent was obtained from all study participants.

### Outcome Variables

Counts of each monocyte subset were defined as the primary outcome variable in the multivariate logistic regression, while AAA disease was defined as the outcome variable in the hazard ratio analysis.

### Clinical Parameters

Serum low-density (LDL) and high-density (HDL) lipoprotein, total cholesterol (TC), triglycerides, non-fasting glucose, hemoglobin (Hb), leukocyte, and C-reactive protein (CRP) concentrations were analyzed in the Institute for Clinical Chemistry and Laboratory Medicine at the TU Dresden using standard laboratory methods. Due to the lack of data on blood chemistry or medical therapies, the number of analyzed patients varies in each group. Cardiovascular risk factors, comorbidities, and prescribed medical therapies were evaluated retrospectively. Hypertension, type 2 diabetes (T2D), heart failure (HF), peripheral artery disease (PAD), and carotid artery stenosis (CAS) were defined by a past documented history of diagnosis or any treatment for these diseases. CAD was defined by a history of myocardial infarction, angina, or treatment for CAD. Smoking was defined as any history of smoking and sex was self-reported.

### Analysis of AAA Diameter and AAA Morphology

The aortic diameter, maximal intraluminal thrombus depth and aortic volumes were determined as described previously.^11^ In brief, aortic diameter was assessed by computed tomography angiography by measuring the distance between the outer adventitia (outer-to-outer-edge) by a single trained observer. Thickness of the ILT was determined in the arterial phase on CT scans following multiplanar reconstruction. The aorta was scanned in an axial position in 1 mm sections. The thickness of the ILT was assessed at the largest distance from the inner surface of the lumen to the outer aortic wall. The AAA volume was measured by two trained people using the automatic segmentation model of the IMPAX EE R20 software (Agfa HealthCare, Mortsel, Belgium). The aorta was scanned with contrast media in the arterial phase in 1 mm sections. The outer wall of the aneurysm and the true lumen were selected manually every 6 mm in the transverse plane. The starting point of the measurement was defined as the cranial end of the aneurysm with a diameter greater than 30 mm. The measurement ended caudal to the dilation or at the aortic bifurcation. Non-recognized areas were cropped manually. The volume of AAA is given in cm^3^. The ILT was included in the total AAA volume measurement but was not specifically tagged for segmentation.

### Blood Samples and Flow Cytometry

Non-fasting venous blood was collected pre-operatively between 2020 and 2024 in 9 mL S-Monovette EDTA (Sarstedt, Nürmbrecht, Germany). Monocyte subpopulations were analyzed using flow cytometry. Cells were stained for 10 min using the following antibody cocktail: CD45-FITC (HI-30, Thermo Fisher Scientific, Darmstadt, Germany), CD16-PerCP710 (CB16, Thermo Fisher Scientific Darmstadt, Germany) and CD14-PE (MφP9, BD Biosciences, Heidelberg, Germnay). Isotype controls were prepared to ensure selectivity of the antibodies. Afterwards, erythrocytes were lysed by the addition of FACS lysing solution (BD Biosciences, Heidelberg, Germnay) for 10 min. Data acquisition was performed on LSR Fortessa (BD Biosciences, Heidelberg, Germnay) and LSR II (BD Biosciences, Heidelberg, Germnay). Data analysis was performed using FlowJo^TM^ Software (Version 10, BD Biosciences, Heidelberg, Germnay). First, debris was excluded by applying SSC and FSC, and events with low FSC and SSC signals were omitted. Before cell aggregates were excluded from the analysis, expression of CD45 against the SSC signal was used to select the monocyte population for further analysis. Finally, classical (CD14++CD16−), non-classical (CD14+CD16++) and intermediate (CD14++CD16+) monocytes were determined according to their expression of CD14 and CD16 **(Supplementary Figure 1)**.

### Statistics

Graph Pad Prism 10.0 (GraphPad Software, Inc., La Jolla, CA, USA) and R Core Team (2021) R: A Language and Environment for Statistical Computing software (R Foundation for Statistical Computing, Vienna/Austria) were used for statistical analysis. A *P*-value ≤0.05 was considered significant. Outliers were tested with the Grubbs outlier test and normality distribution with the D’Agostino and Pearson test. According to the result of the normal distribution test, Mann–Whitney U or unpaired t-test were used to compare AAA and varicose vein patients. Data are shown as box plots with individual values or as scatter dot plots, as indicated in the figure legends. Correlational analysis was done using Pearson correlation (r_P_). Weighted linear regression was performed for variable selection, to assess the influence of age, sex, smoking, hypertension, T2D, CAD, PAD, CRP, HDL-, LDL cholesterol, triglyceride, hemoglobin, leukocyte count, AAA diameter, thickness of the ILT and prescription of insulin, diuretics, statins, ACE, CCB, ARBs, β-blocker, ASA and therapeutic anticoagulation. As some comorbidities strongly influence each other, only age and sex were included in the final analysis. In addition, PAD was included in the multivariate analysis as patients already served as a control group in numerous studies.^8^ Data were analyzed by multivariate logistic regression using each of the monocyte subgroups as a indepedent variable for the presence of AAA disease. Subgroup concentrations were log transformed, and the effect of risk factors was compared with patients in whom the risk factor was not present.

## Results

### Patient Characteristics

The comparison of AAA and venous vein patient’s reveaeled that the AAA group was significantly older and comprised more male patients than varicose vein controls. As expected, both groups differed in their prevalence of cardiovascular risk factors and prescribed pharmacotherapy. A detailed list of risk factors and pharmacotherapy is given in **Table 1**.

**Table 1:** Clinical Characteristics in Patients with Abdominal Aortic Aneurysm (AAA) and Venous Vein varicose. Blood was taken in the non-fasted state. Reference values for the corresponding blood parameters are given in the table. Reference values for LDL and total cholesterol (TC) are based on the European Society of Cardiology (ESC) recommendations. Reference values for fasting glucose were excluded due to the withdrawal in the non-fasted state. **Statistics:** All data are presented as median with minimum and maximum values. Comparison of continuous data was done using the Mann-Whitney U test or t-test. Discrete data were analyzed by Chi-Square test. For t test (a) t-statistic is reported, Mann-Whitney (b) U statistics and Chi-square (c) *X*^2^ is reported. **Abbreviations:** ACE, angiotensin-converting enzyme; ARBs, angiotensin receptor blocker; BMI, body mass index; ASA, acetylsalicylic acid; CAD, coronary artery disease; CCB, calcium channel blocker; HDL, high-density lipoprotein; LDL, low-density lipoprotein; PAD, peripheral artery disease; T2D, type 2 diabetes mellitus; TIA, transient ischemic attack.

| Observational study |  |  |  |  |
| --- | --- | --- | --- | --- |
|  | Varicosis | eAAA | P-value | Statistics |
| **Baseline demographics** |  |  |  |  |
| n included | 33 | 33 |  |  |
| Age – years, mean ± SD | 62.79 ± 8.15 | 70.39 ± 9.41 | <0.001<sup>a</sup> | t=3.51 |
| Sex, male:female, % male | 20:13, 61 | 30:3, 91 | 0.0041<sup>c</sup> | $X^2=8.25$ |
| **Hematology** |  |  |  |  |
| Hemoglobin – mmol/L, mean<br>± SD, n | 8.99 ± 0.73<br>32 | 9.01 ± 1.10<br>33 | 0.948<sup>a</sup> | t=0.07 |
| Reference values 8.60 – 12.10 mmol/L |  |  |  |  |
| Leukocytes – GPt/L, mean ±<br>SD, n | 6.83 ± 1.48<br>32 | 7.64 ± 1.68<br>33 | 0.046<sup>a</sup> | t=2.04 |
| Reference values 3.8 – 9.8 GPt/L |  |  |  |  |
| **Cardiovascular risk factors** |  |  |  |  |
| LDL cholesterol – mmol/L,<br>mean ± SD, n | 4.18 ± 1.50<br>31 | 3.28 ± 1.36<br>33 | 0.014<sup>a</sup> | t=2.53 |
| Reference values <1.40 mmol/L for people with very-high risk |  |  |  |  |
| HDL cholesterol – mmol/L,<br>median with range, n | 1.63<br>(1.00 – 2.62)<br>31 | 1.28<br>(0.75 – 1.98)<br>33 | 0.015<sup>b</sup> | U=331.00 |
| Reference values >0.90 mmol/L for men and >1.10 mmol/L for women |  |  |  |  |
| Total cholesterol – mmol/L,<br>mean ± SD, n | 4.57 ± 1.37<br>31 | 3.52 ± 1.55<br>33 | 0.006<sup>a</sup> | t=0.85 |
| Reference values <4.00 mmol/L |  |  |  |  |
| Triglycerides – mmol/L,<br>median with range, n | 1.18<br>(0.43 – 3.88)<br>31 | 1.20<br>(0.52 – 3.59)<br>32 | 0.582<sup>b</sup> | U=455.50 |
| Reference values 0.35 – 1.70 mmol/L |  |  |  |  |
| Blood glucose – mmol/L,<br>median with range, n | 5.37<br>(4.30 – 6.81)<br>28 | 5.94<br>(4.69 – 11.33)<br>30 | 0.004<sup>b</sup> | U=238.50 |
| Reference value only for fasting glucose possible |  |  |  |  |
| CRP – mg/L, median with<br>range, n | 1.30<br>(0.50 – 21.30)<br>31 | 2.65<br>(0.60 – 16.80)<br>31 | 0.049<sup>b</sup> | U=353.50 |
| Reference values <5.0 mg/L |  |  |  |  |
| Smoking, yes:no – % | 6:27, 18 | 23:10, 70 | <0.001<sup>c</sup> | X<sup>2</sup>=17.78 |
| Hypertension, yes:no – % | 11:22, 33 | 30:3, 91 | <0.001<sup>c</sup> | X<sup>2</sup>=23.24 |
| CAD, yes:no – % | 0:33, 0 | 10:23, 30 | 0.001<sup>c</sup> | X<sup>2</sup>=11.79 |
| HF, yes:no – % | 1:32, 3 | 10:23, 30 | 0.003<sup>c</sup> | X<sup>2</sup>=8.84 |
| CAS, yes:no – % | 0:33, 0 | 7:26, 21 | 0.005<sup>c</sup> | X<sup>2</sup>=7.83 |
| PAD, yes:no – % | 0:33, 0 | 8:25, 24 | 0.003<sup>c</sup> | X<sup>2</sup>=9.10 |
| T2D, yes:no – % | 0:33, 0 | 11:22, 33 | <0.001<sup>c</sup> | X<sup>2</sup>=13.20 |
| BMI – kg/m<sup>2</sup>, median with range, n | 27.20<br>(21.20 – 40.70)<br>33 | 27.25<br>(20.30 – 38.01)<br>33 | 0.677<sup>b</sup> | U=511.50 |
| **Pharmacological therapies** |  |  |  |  |
| Statins, yes:no – % | 3:29, 9 | 24:9, 73 | <0.001<sup>c</sup> | X<sup>2</sup>=26.85 |
| ACE inhibitors, yes:no – % | 3:29, 9 | 15:18, 45 | 0.001<sup>c</sup> | X<sup>2</sup>=10.56 |
| ARB, yes:no – % | 4:28, 13 | 13:20, 39 | 0.014<sup>c</sup> | X<sup>2</sup>=6.08 |
| CCB, yes:no – % | 2:30, 6 | 13:20, 39 | 0.002<sup>c</sup> | X<sup>2</sup>=10.05 |
| ASA, yes:no – % | 2:30, 6 | 25:8, 76 | <0.001<sup>c</sup> | X<sup>2</sup>=32.32 |
| b-blocker, yes:no – % | 4:28, 13 | 17:16, 52 | <0.001<sup>c</sup> | X<sup>2</sup>=11.31 |
| Anticoagulation, yes:no – % | 6:26, 19 | 3:30, 9 | 0.260<sup>c</sup> | X<sup>2</sup>=1.27 |
| Antiplatelet, yes:no – % | 1:31, 3 | 3:30, 9 | 0.317<sup>c</sup> | X<sup>2</sup>=1.00 |
| Diuretics, yes:no – % | 4:28, 13 | 9:24, 27 | 0.137<sup>c</sup> | X<sup>2</sup>=1.49 |
| T2D treatment, yes:no – % | 0:32, 0 | 11:22, 33 | <0.001<sup>c</sup> | X<sup>2</sup>=12.84 |
| Insulin, yes:no – % | 0:32, 0 | 2:31, 6 | 0.157<sup>c</sup> | X<sup>2</sup>=2.00 |

### Phenotypic Heterogeneity of Monocyte Subsets in AAA Patients

In the present study, significants shifts in monocyte subsets were observed in AAA patients compared to controls with varicose veins. In patients with an AAA, there were 1.6-fold lower (*P*=0.001) counts for classical monocytes and 1.9-fold (*P*=0.025) and 1.8-fold (*P*=0.019) higher counts for non-classical monocytes and intermediates, respectively (**Figure 1**). To determine which of the patients’ risk factors and comorbidities influenced the subgroups the most, we performed a linear weighted regression. We found that classical monocytes are relatively robust, since they are barely affected by any of the investigated comorbidities and risk factors **(Supplementary Table 1).** In non-classical monocytes, PAD was associated with an increase, and T2D as well as hypertension were associated with a decrease in this subset **(Supplementary Table 2).** Intermediate monocytes were strongly influenced by the presence of T2D, while sex and hypertension were associated with a decrease in this subset **(Supplementary Table 3).** All the investigated comorbidities and risk factors were seen to highly confound each other. We therefore performed a logistic regression analysis and chose age, sex and PAD for adjustment to analyze whether these factors influence the observed differences between varicose veins and AAA. After adjusting for age, sex and PAD the observed decrease in classical monocytes (*P*=0.001) and increase in non-classical monocytes (*P*=0.040) in AAA patients remained significant (**Table 2**).

**Figure 1:**
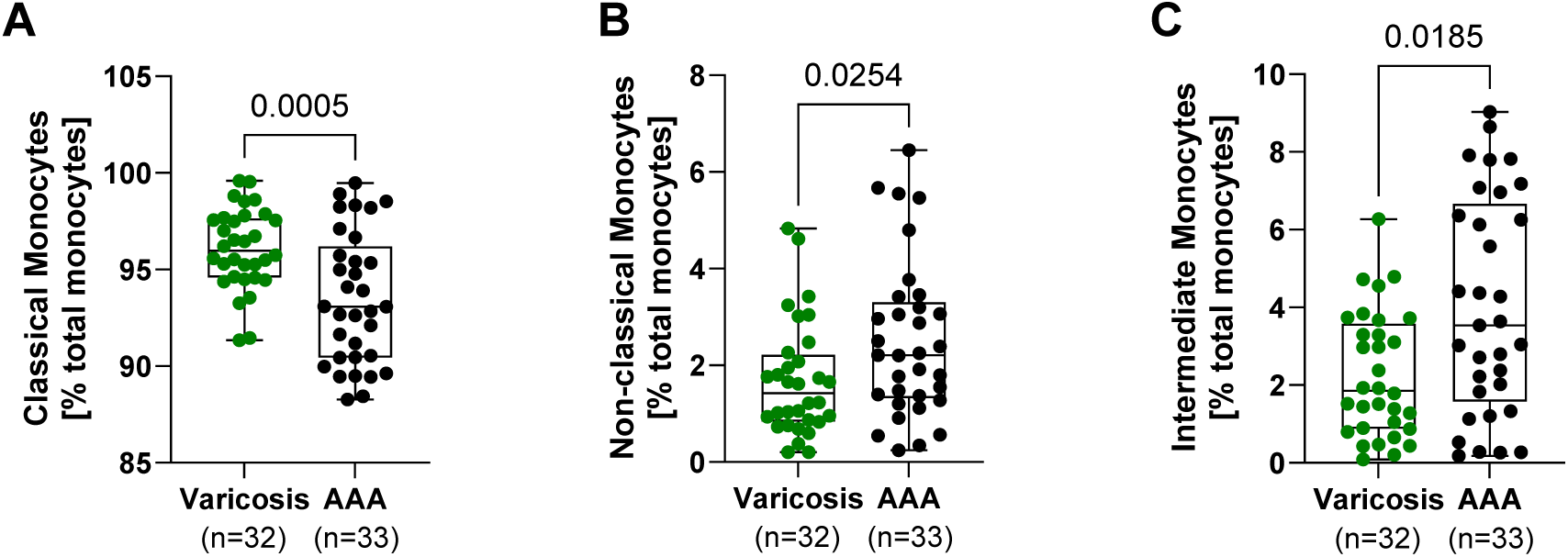
Monocyte Subsets in Patients with AAA and Varicose Vein Controls. Non-fasting venous-blood was collected and the relative frequency of the individual monocyte subpopulations was determined by flow cytometry. **A,** classical (CD14++CD16−), **B,** non-classical (CD14+CD16++) and **C,** intermediate (CD14++CD16+) monocytes. All data are shown as boxplots with individual values. Outliers were identified using the Grubb’s outlier test and one patient in each group was excluded from all further analysis. **A,** Unpaired t-test, **B-C,** Mann-Whitney U test.

**Table 2:** Differences in Monocyte Subsets after Adjusting to Age, Sex and PAD. Monocyte subsets were analyzed using flow cytometry and monocytes are given in % of total monocytes. Data were analyzed by multivariate logistic regression using each of the monocyte subgroups as a diagnostic variable for the presence of AAA disease. The monocyte subgroups were logarithmically transformed, and data were analyzed by multivariate logistic regression using the corresponding subset as a predictor variable. The effects of each risk factor were compared with patients in whom the risk factor was not present (ref=none). The odds ratio (OR) refers to the relative increase in risk of AAA disease as the percentage of the subgroup increases on the logarithmic scale. For sex, ORs refer to men and reference values refer to women. AAA diameter and age show the increase per unit (year and mm, respectively). Unadjusted values were obtained by pairwise comparison of each variable listed in the table with the outcome of having an AAA. Adjusted values were analyzed by holding the effects of the other variables constant and assuming an increase in the monocyte subset on the logarithmic scale. **Abbreviations:** OR, odds ratio.

| Variable | Unadjusted OR<br>[95% CI] | P-value | Adjusted OR<br>[95% CI] | P-value |
| --- | --- | --- | --- | --- |
| **classical monocytes [log10]** |  |  |  |  |
| Group<br>(ref=varicosis) | 0.973 [0.958, 0.987] | <0.001 | 0.968 [0.949, 0.987] | 0.001 |
| Age (years) | 0.999 [0.998, 1.000] | 0.076 | 1.000 [0.999, 1.001] | 0.950 |
| Sex (ref=female) | 1.001 [0.982, 1.021] | 0.895 | 1.014 [0.994, 1.035] | 0.160 |
| PAD (ref=none) | 0.987 [0.963, 1.012] | 0.308 | 1.004 [0.980, 1.029] | 0.754 |
| **non-classical monocytes [log10]** |  |  |  |  |
| Group<br>(ref=varicosis) | 1.504 [1.019, 2.220] | 0.040 | 1.760 [1.058, 2.928] | 0.030 |
| Age (years) | 1.002 [0.981, 1.024] | 0.830 | 0.988 [0.964, 1.012] | 0.331 |
| Sex (ref=female) | 1.024 [0.635, 1.652] | 0.921 | 0.784 [0.462, 1.330] | 0.360 |
| PAD (ref=none) | 1.283 [0.697, 2.360] | 0.417 | 1.026 [0.537, 1.960] | 0.937 |
| **intermediate Monocytes [log10]** |  |  |  |  |
| Group<br>(ref=varicosis) | 1.674 [0.988, 2.836] | 0.055 | 1.940 [0.987, 3.814] | 0.054 |
| Age (years) | 1.024 [0.996, 1.053] | 0.091 | 1.007 [0.975, 1.040] | 0.680 |
| Sex (ref=female) | 0.709 [0.375, 1.343] | 0.287 | 0.563 [0.279, 1.136] | 0.107 |
| PAD (ref=none) | 1.203 [0.527, 2.745] | 0.656 | 0.857 [0.363, 2.024] | 0.721 |

### Monocyte Subsets, AAA Risk and Associations with AAA Morphology

Hazard ratios were calculated to test whether differences in monocyte subgroups lead to an increased risk of AAA disease. A reduced number of classical monocytes was associated with an increased risk of AAA (*P*=0.002). Furthermore, increased levels of non-classical (*P*=0.030) and intermediate monocytes (*P*=0.006) led to a higher risk for AAA disease (**Table 3**).

**Table 3:**
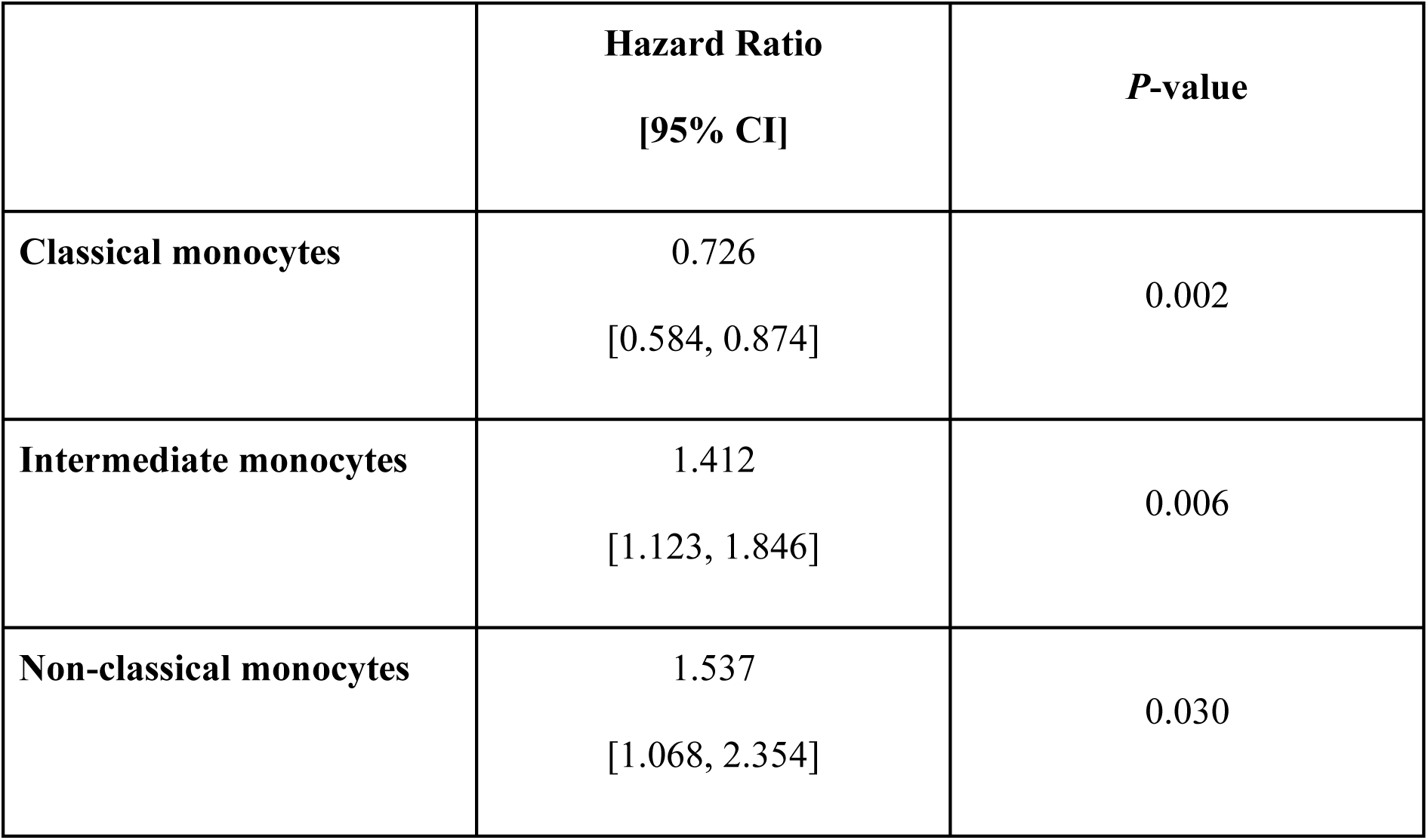
Hazard Ratios of Monocyte Subsets and AAA risk. Monocyte subsets were analyzed using flow cytometry and monocytes are given in % of total monocytes. Logistic regression was used to obtain hazard ratio of incident AAA per one unit increase in monocytes subsets on log scale.

|  | **Hazard Ratio**<br>**[95% CI]** | ***P*-value** |
| --- | --- | --- |
| **Classical monocytes** | 0.726<br>[0.584, 0.874] | 0.002 |
| **Intermediate monocytes** | 1.412<br>[1.123, 1.846] | 0.006 |
| **Non-classical monocytes** | 1.537<br>[1.068, 2.354] | 0.030 |

### Monocytes and Correlations with AAA Morphology

A correlational analysis was performed to test whether the different monocyte subgroups correlate with the morphology of the AAA and can therefore serve as surrogate markers. Classical monocytes showed a trend to increase with AAA diameter (**Figure 2A**), while no correlations with AAA volume or ILT thickness were found (**Table 4**). Conversely, non-classical monocytes decreased with increasing AAA diameter (r_P_=-0.64, *P*<0.001) and increasing AAA volume (r_P_=-0.50, *P*=0.003) (**Figure 2B,C**). Intermediate monocytes showed a trend towards a positive association with maximum depth of the ILT (r_P_=0.32, *P*=0.067) (**Figure 2D**). **Table 4** provides an extended overview of the tested correlations.

**Figure 2:**
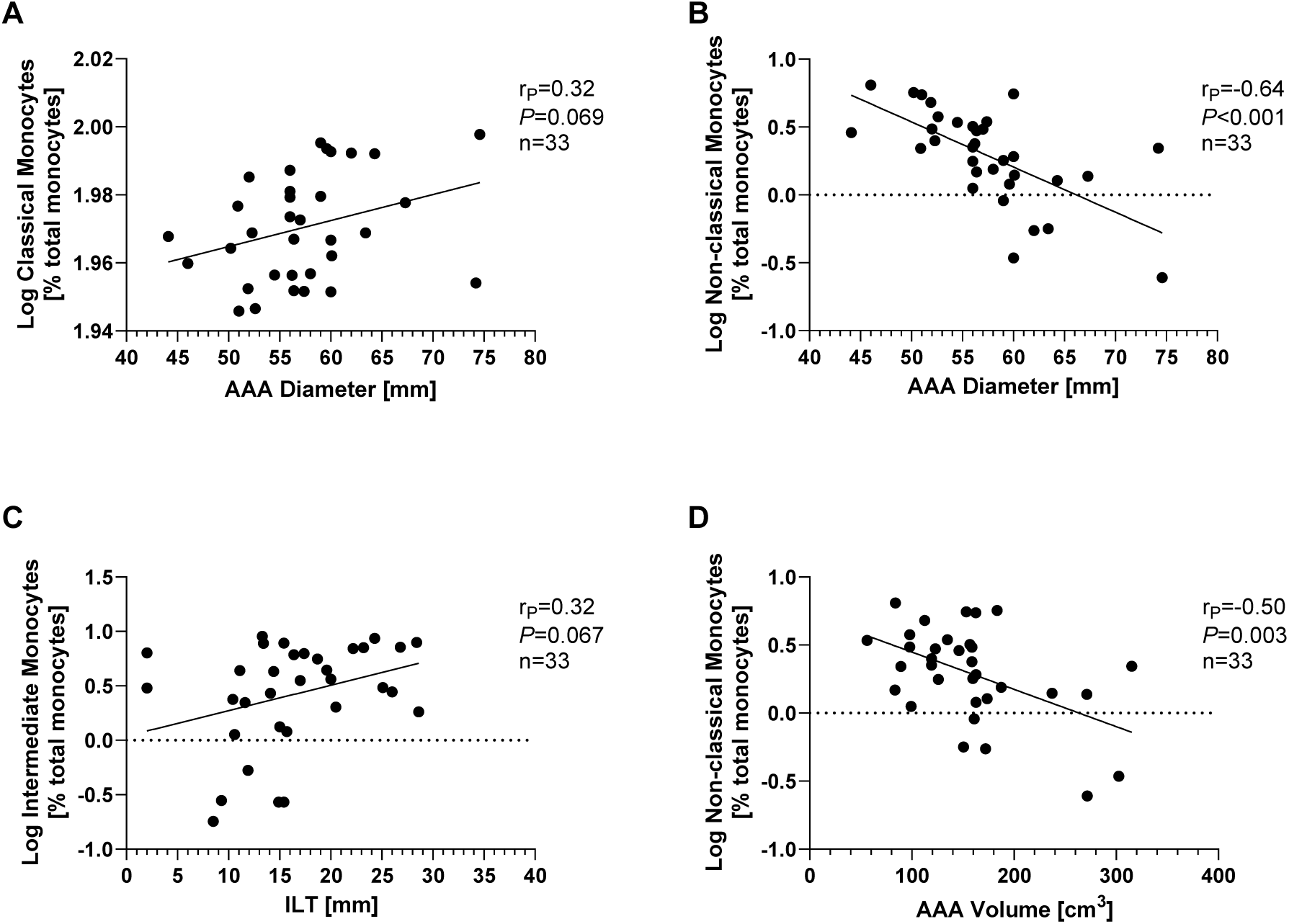
Correlation of Monocyte Subsets and AAA Morphology. Monocyte subsets were analyzed using flow cytometry and are given as % of total monocytes. The data were logarithmically transformed and analyzed for correlations with the morphology of the AAA. Correlation of **A,** classical monocytes with AAA diameter, **B,** AAA diameter and non-classical monocytes, **C,** non-classical monocytes and AAA volume and **D,** ILT and intermediate monocytes. **Statistics:** Grubb’s outlier test was used to identify outliers. One AAA patient was identified as an outlier and excluded from all further analysis. Correlational analysis was done using the Pearson correlation. r_P_, Pearson rank correlation coefficient.

**Table 4:** Summary of Correlations Between Monocyte Subsets and AAA Morphology. Monocyte subsets were analyzed using flow cytometry and monocyte subsets are given in % of total monocytes. Data were log transformed. Correlational analysis was performed using Pearson correlation. Outliers were identified using Grubb’s outlier test and excluded from further analysis. **Abbreviations:** AAA, Abdominal aortic aneurysm, **ILT**, Intraluminal thrombus, **r_P_,** Pearson’s rank correlation coefficient.

|  | classical monocytes |  | non-classical monocytes |  | intermediate monocytes |  |
| --- | --- | --- | --- | --- | --- | --- |
|  | rp | P-value | rp | P-value | rp | P-value |
| **AAA diameter** | 0.32 | 0.069 | **-0.64** | **<0.001** | -0.20 | 0.265 |
| **ILT** | -0.21 | 0.23 | 0.19 | 0.298 | **0.32** | **0.067** |
| **AAA volume** | 0.27 | 0.13 | **-0.50** | **0.003** | -0.13 | 0.481 |

## Discussion

Monocytes are important inflammatory mediators in cardiovascular disease and also in AAA as a particular CVD. Depending on the expression of their surface markers, monocytes are classified into different subgroups that have different functions. Knowledge about their expression patterns may contribute to the understanding of specific disease mechanisms as these cells migrate into inflamed tissue and differentiate into macrophages. Moreover, this would allow the identification of patients at high risk and provide useful surrogate markers for AAA morphology. Although an increase in intermediate subsets has been demonstrated in AAA^8^, nothing is known whether these subsets correlate with AAA diameter, AAA volume and intraluminal thrombus as main morphologic features of AAA. Determination of surrogate markers that correlate with the maximum AAA diameter is of great interest, as the AAA diameter is the most important marker for decisions on surgical therapy.^12^ Furthermore, there is no information on whether any of these subgroups can predict the risk of having AAA.

The present study showed a lower count of classical monocytes (CD14++CD16−) and higher counts of intermediate (CD14++CD16+) and non-classical monocytes (CD14+CD16++) in patients with AAA. Our data are, at least in part, supported by previously published studies.^7,8^ The decreased proportion of classical monocytes could be explained by findings in mice after induction of acute myocardial infarction. In the first phase, CD14+CD16− monocytes accumulate early at the site of injury to clear necrotic debris and promote healing of the damaged tissue. CD14+CD16++ dominate in the second phase and promote healing through the accumulation of myofibroblasts, angiogenesis and collagen deposition.^13^ The data herein eventually underline an imbalance between both subsets in patients with AAA towards intermediate subsets that promote fibrotic remodeling in the AAA wall. While non-classical monocytes remain in the bloodstream, classical monocytes could transmigrate through the endothelium into the vessel wall.^14^ This could be a possible explanation for the depletion of the classic monocytes, which could be due to an increased migration into the AAA.

Non-classical monocytes are the most inflammatory subset but also show patrolling function.^15^ Studies in patients with CAD^16^ and subclinical atherosclerosis demonstrated increased non-classical subsets^17^ and therefore coincidence the data of the present study. Herein, a higher count of non-classical was found in AAA, which is different to data previously published by Klopf et al.^8^ Most often, patients with PAD served as controls in biomarker studies in AAA.^18^ PAD is one of the most common initial manifestations of T2D^19^ and T2D itself is inversely associated with AAA disease.^20^ It might be of speculation that a confounding of protective and adverse effects could occur.

To further understand the link between AAA risk and blood monocytes, we analyzed whether a specific subtype of monocyte was associated with increased risk of having AAA disease. We were able to show that a lower proportion of classical monocytes was associated with a higher risk of AAA, while an increase in non-classical and intermediate monocytes was associated with an increased risk of AAA. It could be that the increase in intermediate and non-classical subsets occurs at the expense of classical monocytes. Similar results were demonstrated in patients with T2D.^21^ Intermediate monocytes have been demonstrated to be an independent predictor of cardiovascular events in patients with atherosclerotic diseases.^22^ Similar results were obtained for non-classical monocytes.^22^ The increased risk might be explained by the proinflammatory role of intermediate and non-classical monocytes.^22^ Interestingly, the percentage of classical monocytes might be a protective factor for major adverse outcomes in coronary artery disease.^23^ Classical monocytes are known to be phagocytic but have no inflammatory potential.^15^ Given their protective role, the decrease in AAA and their association with AAA risk might be a hint to a loss of their function.

The higher percentage of non-classical, CD14+CD16++ monocytes was inversely associated with the AAA diameter and AAA volume. Analyzing the AAA volume has the advantage of including the morphology of the aneurysm.^24^ AAA volume can be useful when the diameter fails or for monitoring saccular AAA, since there is a weaker correlation between increased diameter and rupture risk in this type of AAA.^25^ Finally, AAA volume might be a better predictor for AAA growth.^26^ A negative correlation between non-classical, CD14+CD16++ monocytes with the intima-media-thickness was shown in patients with carotid artery stenosis^27^ and in peripheral artery disease where non-classical subsets decreased with the severity^5^. CD14+CD16++ monocytes (or Ly-6C^lo^ in mice) are involved in healing of the ischemic myocardium and they initiate the differentiation towards M2 macrophages, known to be involved in tissue repair. It might be of speculation, but the inverse relationship could be explained by temporal patterns and lack of their protective and healing functions^27^, although they are higher in patients with AAA. Another possibility that could explain the inverse relationship between AAA diameter and non-classical monocytes is that non-classical monocytes migrate into the AAA, i.e. as the diameter increases, more cells migrate in and their concentration in the blood decreases. In addition, the patients examined here are in the final stage of the disease and exhibit the typical degeneration of the AAA wall.^18,28^

The majority of AAA are covered by an intraluminal thrombus (ILT). The ILT contains fibrin, inflammatory cells, platelets and red blood cells and affects growth and rupture of the AAA.^29,30^ Herein, intermediate (CD14++CD16+) subsets tended to correlate with the thickness of the ILT in AAA-patients. Intermediate monocytes are capable of antigen presentation, secreting pro-inflammatory cytokines and to regulate immune responses.^31^ It has been demonstrated that intermediate monocytes are present in the ILT but in lower amounts than in peripheral blood of AAA patients.^32^ The positive association could reflect the inflammatory nature of the ILT or also cells that are released by the ILT.

Overall, the present study showed age- and sex-independent shifts in classical, non-classical and intermediate subsets in AAA, suggesting disease-specific mechanisms. An decrease in classical monocytes and the increases in intermediate and non-classical monocytes were associated with risk for having AAA. The number of non-classical monocytes correlated inversely with AAA diameter and AAA volume and could therefore be a useful surrogate marker for AAA morphology, beyond the AAA diameter. Especially the lowering in phagocytic classical monocytes and the increase in non-classical monocytes should be considered in future studies.

### Limitations

The present study is a descriptive, observational study on a very small cohort of patients. Varicose vein patients were used as a control group and their limitation is their different comorbidity and risk factor profile. The relativley small number of patients included and the large number of confounding factors may have influenced the results. Analyzing patients in whom the risk factors have been adjusted prior, e.g. in a case-control study, could minimize these effects. Furthermore, the results obtained here do not allow to establish a causal relationship between the different monocyte subsets and the development of AAA which is due to the study design. Finally, the main discrepancy when comparing data from flow cytometry with the present study is the pre-selection of cells, the declaration of subgroups based on the expression of CD14 and CD16 and the gating strategy in flow cytometry.

## Data Availability

All data are published within the manuscript.

## Acknowledgement

Flow cytometry measurement and analysis was done at the Flow Cytometry Core Facility at TU Dresden, University of Technology. We would like to thank Dr Anne Gompf, Head of the Flow Cytometry Core Facility, for her excellent technical experience.

## Non-standard abbreviations and acronyms

AAA: abdominal aortic aneurysm
ACE: angiotensin-converting enzyme
ARBs: angiotensin receptor blocker
ASA: acetylsalicylic acid
BMI: body mass index
CAD: coronary artery disease
CAS: carotid artery stenosis
CCB: calcium channel blocker
CVD: cardiovascular disease
Hb: hemoglobin
HDL: high-density lipoprotein cholesterol
HF: heart failure
LDL: low-density lipoprotein cholesterol
PAD: peripheral artery disease
T2D: Type 2 diabetes mellitus
TC: total cholesterol

## Notes

### Competing Interest Statement

The authors have declared no competing interest.

### Clinical Trial

The study is a basic science study.

### Funding Statement

No external funding was received.

### Author Declarations

The study was approved by the ethics committee of the Technische Universität Dresden (EK 151042017).

